# Safety and immunogenicity of recombinant hepatitis E vaccine in healthy pregnant women between 14 and 34 weeks of gestation and non-pregnant women of reproductive age: Protocol for a Phase II, randomized, observer-blinded, placebo-controlled trial

**DOI:** 10.64898/2026.08.07.26359927

**Authors:** Katerina Rok Song, Imran Nisar, Jiyoung Lee, Lei Yang, Deok Ryun Kim, Afida Riskiana, Nigus Telele, Aneeta Hotwani, Nadia Ansari, Sidhar Nausheen, Lumaan Sheikh, Wei Chen, Xiaoshan Yu, Rui Wang, Madeleine Blunt, Kawsar R. Talaat, Brittany L. Kmush, Fyezah Jehan, Julia Lynch

## Abstract

**Introduction:** Hepatitis E virus (HEV) in pregnancy is associated with high maternal and perinatal morbidity and mortality. The safety and efficacy of the recombinant protein HEV vaccine (HEV239, Hecolin^®^) have been established in non-pregnant adult populations but there is limited information among pregnant women. This trial has two co-primary objectives: 1) to assess pregnancy-related and/or serious safety events among pregnant women between 14 and 34 weeks of gestation receiving two Hecolin^®^ doses four weeks apart compared to placebo recipients, and 2) to determine immune non-inferiority of pregnant recipients of two Hecolin^®^ doses four weeks apart compared to non-pregnant women.

**Methods and Analysis:** This is a multi-site, randomized, observer-blinded, placebo-controlled vaccine safety and immunogenicity trial in pregnant women and non-pregnant women of reproductive age in Karachi, Pakistan. A total of 2,358 healthy women will be enrolled, including 2,208 pregnant women between 14 and 34 weeks of gestation, who will be randomized in a 1:1 ratio (stratified by gestational age, 14–27 and 28–34 weeks) to receive either Hecolin^®^ or a normal saline placebo in two doses administered 1 month apart during pregnancy and a third dose administered postpartum, approximately 5 months after the second dose. A third arm of 150 non-pregnant women aged 16–45 years will receive Hecolin^®^ on 0, 1, and 6 months. The co-primary outcomes will be (i) the proportion of pregnancy-related AESIs and SAEs in pregnant participants from the first dose until the end of study follow-up, compared with placebo, and (ii) the geometric mean concentration (GMC) of anti-HEV IgG at four weeks after the second dose, comparing pregnant vaccine recipients with non-pregnant vaccine recipients (non-inferiority margin of 0.67 for the GMC ratio). Immunogenicity will be evaluated in a pre-specified subset of 300 participants receiving Hecolin^®^, including 150 pregnant participants and 150 non-pregnant participants. Secondary outcomes will include maternal, neonatal, and infant safety outcomes, as well as immunogenicity according to the number of Hecolin^®^ doses received and the trimester of vaccination.

**Ethics and Dissemination:** The trial was approved by the National Bioethics Committee (NBC) of Pakistan (Reference number: 4-87/NBC-910), the institutional Ethics Review Committee (ERC) of the Aga Khan University (Reference number: 8298), and the Institutional Review Board (IRB) of the International Vaccine Institute (IVI) (Reference number: 2022-007). All participants will provide written informed consent in accordance with Good Clinical Practice. The results will be submitted to World Health Organization (WHO) Strategic Advisory Group of Experts in Immunization (SAGE), and disseminated through conference presentations, and peer-reviewed publications.

**Article Summary:**

- This study will directly enroll pregnant women, the population at highest risk of HEV-related maternal and perinatal morbidity and mortality, and will employ a randomized, observer-blinded, placebo-controlled clinical trial design to minimize bias in outcome assessment.
- The study will be conducted in peri-urban Karachi, Pakistan, an area endemic for hepatitis E virus (HEV) genotype 1 with documented outbreaks and ongoing endemic transmission, making the findings directly relevant to similarly HEV-endemic settings. While the findings are likely generalizable to other HEV-endemic settings, the high background burden of pregnancy-related adverse outcomes in Pakistan, including preterm birth, low birth weight, and maternal, fetal, and infant mortality, may result in higher observed adverse event rates in this study population.
- Participants in the first trimester of pregnancy were excluded; therefore, the safety of vaccine exposure during early gestation will not be assessed.
- Follow-up will be limited to six months postpartum, precluding assessment of the long-term durability of the immune response.
- A minimum interval of at least two weeks will be required between administration of the investigational product and other vaccines routinely given in pregnancy (e.g., tetanus toxoid), to avoid potential immune interference with immunogenicity assessment. Therefore, the safety and immunogenicity of concomitant administration with routine maternal vaccines will not be evaluated in this trial.

## Introduction

Hepatitis E virus (HEV) infection is one of the most common causes of acute viral hepatitis globally, with an estimated 19.47 million infections annually in 2021. [1,2] HEV is transmitted to humans through contaminated water and other fecal–oral routes, predominantly for genotypes 1 and 2, and through zoonotic transmission via infected pork products or direct animal contact for genotypes 3, 4, and 7. [3–5] Multiple HEV outbreaks have been documented, particularly in Asia and Africa where most have occurred in displaced persons camps. [2,5–7] HEV infection is usually self-limited in healthy adults and may be asymptomatic but can cause severe disease in specific populations. In pregnant women, acute HEV infection is associated with high rates of maternal mortality (5-31%) and complications including intrauterine growth restriction, preterm birth, maternal fulminant hepatic failure, and fetal demise. [2,8,9]

HEV is a vaccine-preventable disease, and Hecolin^®^ has been available since 2011 and is licensed in China, India, and Pakistan. [10,11] Hecolin^®^ is a recombinant protein vaccine based on a fragment of the HEV genotype 1 ORF2 capsid protein expressed in *Escherichia coli*. [10–12] Across Phase I–III clinical trials, Hecolin^®^ administered as a three-dose series (0, 1, and 6 months) has demonstrated a favorable safety profile and strong immunogenicity. In the pivotal Phase III randomized controlled trial and its long-term follow-up, the vaccine demonstrated high efficacy, with per-protocol efficacy of 100% (95% CI: 72.1–100) at 19 months, 93.3% (95% CI: 78.6–97.9) at 4.5 years, and 86.6% (95% CI: 73.0–94.1) at 10 years after immunization. [13–15] In 2015, the World Health Organization (WHO) recommended Hecolin^®^ be considered for use where the risk of hepatitis E is high. WHO concluded that evidence on the safety, immunogenicity, and efficacy of Hecolin^®^ during pregnancy was insufficient to recommend routine vaccination in pregnant women. However, they acknowledged that there may be special situations, such as during active outbreaks, where use of the vaccine should be considered to mitigate consequences in high-risk groups, such as pregnant women. [16] Following a review of new evidence, the WHO Strategic Advisory Group of Experts (SAGE) on Immunization updated its recommendations in 2024, concluding that in fragile, conflict-affected, and vulnerable settings with documented HEV circulation, the benefits of vaccinating women of childbearing age (≥16 years), including pregnant women, outweigh the potential risks. SAGE also highlighted the need for additional evidence on the safety and effectiveness of Hecolin^®^ during pregnancy to further inform policy recommendations.

Safety data on Hecolin^®^ use during pregnancy are generally reassuring, although more recent data have yielded a mixed picture. Pregnancy safety data are available from two post hoc analyses of manufacturer-sponsored trials conducted by Innovax. These include a Phase III trial of Hecolin^®^, which compared Hecolin^®^ with a hepatitis B vaccine, and a Phase III trial of Cecolin (a bivalent HPV vaccine), in which Hecolin^®^ was used as the control. Both trials evaluated inadvertent vaccination during pregnancy (n=68 in the Hecolin^®^ Phase III trial and n=140 in the Cecolin Phase III trial), both of which showed no evidence of increased risk of adverse pregnancy outcomes. [17, 18] Observational data from a mass vaccination campaign in response to an HEV outbreak in an internally displaced persons camp in South Sudan were used as part of an emulated target trial, along with unvaccinated women from the community matched by gestational and maternal age and propensity to be vaccinated. [19] The primary outcomes were composite fetal loss (spontaneous abortion or stillbirth) for Hecolin^®^ exposure any time during pregnancy and spontaneous abortion following first-trimester exposure, assessed by participant interview within 28 days after the expected delivery date and verified where possible. In the primary analysis, composite fetal loss was not significantly different between 1,928 pregnant women receiving at least one dose during pregnancy (7.2%, 95% CI: 5.6–8.7) and unvaccinated controls (6.1%, 95% CI: 3.7–9.2; risk ratio=1.2 (95% CI: 0.7-1.9)). [19] In analysis limited to women less than 90 days gestational age, risk of spontaneous abortion was also not significantly different between vaccinated women and matched unvaccinated controls (10.5% (95% CI: 8.0-14.4) vs. 11.7% (95% CI: 6.4-18.8); risk ratio=0.9 (95% CI: 0.5-1.9)). This study also documented high effective protection (adjusted rate=84%, 95% CI: −208.5-99.2) with two doses of Hecolin^®^ compared to matched unvaccinated community controls. [6,19]

However, a Phase IV cluster-randomized, double-masked trial comparing Hecolin^®^ to hepatitis B virus (HBV) vaccine among women of reproductive age conducted in Bangladesh targeted to enroll non-pregnant women of reproductive age. Among women inadvertently exposed to Hecolin^®^ either during or early pregnancy, the risk of spontaneous abortion was higher than among women receiving HBV vaccine (10.5% (22/209) vs. 5.3% (14/266), adjusted relative risk (aRR)=2.1, 95% CI: 1.1 – 4.1)). Similarly, among women vaccinated within 90 days before the last menstrual period, spontaneous abortion was more frequent in the Hecolin^®^ group (8.0% (32/398) vs. 4.0% (18/453), aRR=1.9, 95% CI:1.1–3.2, p=0·013), whereas the risk of stillbirths and elective terminations did not differ significantly. [20,21] The heterogeneity in study design, pregnancy exposure definitions, outcome measures, and data collection methods highlights a significant knowledge gap, which has led to continued recommendations for further research to clarify Hecolin^®^ safety in pregnancy. [22]

### Rationale for the proposed study

Existing safety data largely come from studies that either excluded pregnant women or discontinue vaccination after pregnancy was identified. [13,20] Consequently, robust data on the safety and immunogenicity of Hecolin^®^ during the second and third trimesters of pregnancy remain limited. Yet, among the more pressing evidence gaps to inform policy on vaccine use is the safety, immunogenicity and effectiveness of Hecolin^®^ among women in their second and third trimesters who have the highest risk for morbidity and mortality from HEV infection. Nesbitt *et al.* documented successful Hecolin^®^ mass vaccination in the context of slowing an outbreak, where morbidity and mortality risks outweighed those of potential vaccine risks in pregnancy. [6,19,23] Moreover, Hecolin^®^ offers great promise to reduce maternal mortality, particularly in hyperendemic settings, such as Bangladesh where the maternal mortality ratio and neonatal mortality rate from HEV are estimated to be 4.7 (95% CI, 1.6–11.4) and 22.5 (95% CI, 10.5–34.5) per 100 000 live births, respectively. [24] Further, a two-dose Hecolin^®^ regimen administered one month apart is operationally feasible during pregnancy, particularly in outbreak responses or endemic settings where antenatal care is often initiated during the second trimester or later. [6,25] Given the substantial burden of HEV-related maternal morbidity and mortality in South Asia and Africa, this trial will evaluate the safety and immunogenicity of a two-dose Hecolin^®^ regimen administered during pregnancy, followed by completion of the third dose postpartum. This schedule is supported by the high short-term protection observed after two doses in previous studies [26,27] and reflects the practical challenges of completing a three-dose schedule during pregnancy. Demonstrating the safety and immunogenicity of Hecolin^®^ in pregnant women could support their inclusion in future vaccination programmes during outbreaks and other high-risk settings.

### Objectives and outcomes

The primary safety objective is to evaluate the pregnancy-related adverse events of special interest (AESIs) and serious adverse events (SAEs) among pregnant recipients of two doses of Hecolin^®^ administered four weeks apart between 14 and 34 weeks of gestation as compared with pregnant placebo (normal saline) recipients. Safety data will be collected using the following approaches to capture all adverse events (AEs): immediate AEs (30 minutes after each injection) assessed and recorded at the study site; solicited AEs (7 days after each injection) recorded in a structured written diary which include solicited injection site and systemic events, and unsolicited AEs, and concomitant medications use, with data collected during daily telephone follow-up and/or the next study visit; unsolicited AEs (28 days after each injection) collected at the next scheduled study visit; and AESI and SAEs reported at any point during the study period. Pregnancy-related AESIs include hypertensive disorders of pregnancy (gestational hypertension, pre-eclampsia, and pre-eclampsia with severe features, including eclampsia), intrauterine growth restriction, gestational diabetes mellitus, pathways to preterm birth (premature preterm rupture of membranes, preterm labor, and medically indicated preterm delivery), chorioamnionitis, antepartum and postpartum hemorrhage, and placental abruption. SAEs will be defined according to established regulatory seriousness criteria, including persistent or significant disability and the need for hospitalization or other medically important healthcare intervention. The primary immunogenicity objective is to demonstrate immune non-inferiority of pregnant recipients compared with non-pregnant recipients following two doses of Hecolin^®^ administered four weeks apart. This outcome will be measured using the geometric mean concentration (GMC) of anti-HEV IgG at four weeks after the second dose.

The secondary safety objective is to characterize the safety profile of Hecolin^®^ in pregnant women and to evaluate neonatal and infant safety among offspring born to Hecolin^®^ recipients compared with placebo recipients.

Secondary maternal safety outcomes will include immediate adverse events, solicited local and systemic adverse events, and unsolicited adverse events. Neonatal and infant safety outcomes will include AESIs, SAEs, and growth and developmental outcomes through six months of age. Neonatal and infant AESIs include congenital anomalies, low birth weight (including very low birth weight and extremely low birth weight), preterm birth, neonatal encephalopathy, hypoxic-ischaemic encephalopathy, birth asphyxia, and sudden infant death syndrome.

The secondary immunogenicity objective is to demonstrate immune non-inferiority of pregnant recipients compared with non-pregnant recipients following one, two and three doses of Hecolin^®^, and to compare immune responses between women vaccinated during the second trimester (14–27 weeks’ gestation) and those vaccinated during the third trimester (28–34 weeks’ gestation).

Secondary immunogenicity outcomes will be measured using anti-HEV IgG geometric mean concentrations (GMCs) and seroconversion rates (SCRs), defined as a ≥4-fold increase in anti-HEV IgG concentration from baseline, measured four weeks after the first, second, and third doses. Comparisons will be performed between pregnant and non-pregnant participants following one, two, and three doses, and between women vaccinated during the second and third trimesters following the second dose.

Exploratory objectives include evaluating the mode of delivery among pregnant participants receiving Hecolin^®^ or placebo, further characterizing maternal and infant anti-HEV antibody responses, including transplacental antibody transfer and the persistence of antibodies in infant serum and breast milk during the first six months after birth, and exploring the impact of baseline anti-HEV serostatus on the safety and immunogenicity of Hecolin^®^.

## Methods and Analysis

### Trial design and setting

This is a Phase II randomized, observer-blinded, placebo-controlled trial evaluating the safety of Hecolin^®^ administered between gestational ages 14 0/7 and 34 6/7 weeks in healthy pregnant women, as well as its immunogenicity, with a non-pregnant comparator arm. The trial is sponsored by the International Vaccine Institute (IVI) and conducted at a single institution, the Aga Khan University (AKU), across multiple recruitment and delivery sites in Karachi, Pakistan.

Participants are recruited from five sites: four AKU-affiliated antenatal referral centers in peri-urban, low-income areas of Karachi (Rehri Goth, Bhains Colony, Ibrahim Hydari, and Ali Akbar Shah) and the AKU Kharadar Hospital for Women and Children. All investigational product administration, blood and breast milk sampling, and infant follow-up are coordinated through the AKU Clinical Trial Unit. Participants are offered the option to deliver at the Kharadar Hospital for Women and Children. All laboratory assays are performed at the AKU Clinical Laboratory and the AKU Infectious Disease Research Laboratory (IDRL).

Hecolin^®^ is licensed for use in Pakistan. The country has reported HEV outbreaks and HEV seroprevalence among pregnant women ranging from 19 to 66% in published studies. [28–31]

During study design, vaccines recommended during pregnancy, such as tetanus-containing vaccines (Td/Tdap) and influenza vaccine, were considered as potential comparators. However, variability in Td/Tdap dosing schedules based on prior vaccination history, as well as the seasonal availability of influenza vaccine, could introduce methodological complexities. Therefore, placebo was selected as the control. Participants in the placebo group will be offered Hecolin^®^ in a routine practice setting after completion of the study and unblinding.

### Recruitment and eligibility

Potential participants are prescreened and recruited through information sessions provided by study staff to pregnant individuals seeking antenatal care (ANC) or non-pregnant women seeking routine health services through the five referral centers, Rehri Goth, Bhains Colony, Ibrahim Hydari, Ali Akbar Shah, and AKU Kharadar Hospital, using Institutional Review Board- and site Ethics committee-approved methods and materials. Self-selecting participants are referred to the AKU Clinical Trial Unit for a screening visit appointment scheduled by study staff present at the affiliated health centers. A total of 2,358 participants will be enrolled, comprising 1,104 pregnant women at 14-27 weeks’ gestation, 1,104 pregnant women at 28-34 weeks’ gestation, and 150 non-pregnant healthy women.

At the screening visit, potential participants will receive study information through a discussion with trained study staff, who will then obtain written informed consent. The informed consent discussion will be conducted in the participant’s preferred language. Witnessed consent with a thumbprint as confirmation of consent will be available for illiterate individuals.

The screening visit assesses eligibility for one of two study populations: pregnant women of gestational age between 14 0/7 and 34 6/7 weeks and 16-45 years of age or non-pregnant women between 16-45 years of age who are using a reliable contraceptive method, if presumed fertile.

The eligibility criteria were developed in accordance with the approved Hecolin^®^ product information and considerations specific to maternal immunization trials. Key inclusion criteria included willingness to comply with study procedures, absence of significant medical conditions that could interfere with study participation, and good general health as determined by medical history, physical examination, and laboratory evaluation. Additional eligibility criteria for pregnant participants include a viable singleton pregnancy and the absence of maternal or fetal conditions associated with a high risk of adverse pregnancy outcomes.

Participants are excluded if they have previously received a hepatitis E vaccine, have a known hypersensitivity to vaccine components, are immunocompromised or receiving immunosuppressive therapy, have recently received blood products or another vaccine within the protocol-defined intervals, or have any clinically significant condition that could compromise participant safety or the evaluation of study outcomes. Pregnant women are additionally excluded if they have current pregnancy complications or obstetric conditions associated with increased maternal or fetal risk, plan pregnancy termination, or have other significant obstetric or gynecologic conditions considered by the investigator to preclude study participation. Temporary exclusion criteria allow enrolment to be deferred until the condition has resolved.

### Study interventions

Eligible participants are randomized through a centralized Interactive Web Response System (IWRS) using undisclosed block sizes. Pregnant women are allocated in a 1:1 ratio to Arm 1 (Hecolin^®^) or Arm 2 (placebo), with equal allocation across the two gestational-age strata. Within each pregnant arm, 150 participants (75 per gestational-age stratum) are enrolled in the immunogenicity subset. Arm 3 comprises 150 eligible non-pregnant women who receive Hecolin^®^, and all participants in this arm constitute the non-pregnant immunogenicity comparator.

Pregnant participants (Arms 1 and 2) receive two doses of the investigational product, four weeks apart, during pregnancy. A third dose is administered postpartum, approximately 20 weeks after the second dose. If the scheduled third-dose interval falls before delivery, the visit (V6) is rescheduled to occur within 14 days after delivery, and subsequent visits are adjusted accordingly to maintain the protocol-defined visit windows. Participants assigned to placebo (Arm 2) will be offered Hecolin^®^ in a routine practice setting following completion of their final study visit and unblinding, to ensure equal access to the vaccine. Non-pregnant participants (Arm 3) receive Hecolin^®^ on the licensed 0, 1, and 6 months schedule.

### Biologic specimen sampling

For the immunogenicity subset, blood samples will be collected before and one month after each investigational product administration. The final maternal blood sample will be collected six months after the third dose. Maternal blood and breast milk samples will also be collected at delivery and at six weeks and six months postpartum.

Eight-milliliter venous blood samples will be collected from pregnant and non-pregnant participants, by trained staff following standard universal precautions. After clotting for 30 minutes at room temperature, blood samples will be centrifuged to separate serum, which will be transferred into cryovials and stored at −80°C until testing. For pregnant participants, 10 mL of breast milk will be collected, either by manual expression or using an electric breast pump, at delivery and the six-week and six-month postpartum visits. For infants born to mothers in the immunogenicity subset, 10 mL of umbilical cord blood will be collected at delivery or, if cord blood is unavailable, 3 mL of infant venous or capillary blood (heel stick) will be collected within seven days of birth. Subsequently, 3 mL of infant venous blood will be collected at the six-week and six-month visits. All samples collected at the clinical sites will be stored at 2-8°C and transported to the AKU laboratory for processing, aliquoting, storage, and subsequent antibody testing according to standard operating procedures.

Both serum and breast milk samples will be tested for anti-HEV IgG and anti-HEV IgM antibodies using the Wantai HEV IgG and Ig M ELISA kits (Beijing Wantai Biological Pharmacy Enterprise, Beijing, China) following manufacturer’s instructions for serum and as described in the study standard operating procedures for breast milk samples. Anti-HEV IgG seropositive samples will be further tested using an in-house quantitative ELISA based on the WANTAI HEV IgG ELISA kit and WHO Reference Reagent for antibodies to hepatitis E virus, human serum (NIBSC code: 95/584). [32] The anti-HEV IgG antibody concentration will be reported in international units per milliliter (IU/mL).

Biologic specimens from participants providing additional informed consent for future use of human biological specimens will be stored at the IVI Immunology Laboratory using coded identifiers that permit linkage of biologic specimens with phenotypic and clinical data while maintaining participant confidentiality and study masking. Stored samples may be used for additional assessment of immunogenicity, study of possible immune correlates of protection, validation of assays, testing of new assays, and other safety-related research, depending on the consent status of the samples for future research. Serum samples may be pooled with others for future research and may be used in the development of an International Reference Reagent for antibodies to hepatitis E virus by WHO or other organizations.

### Safety considerations, suspending investigational product use, and oversight

Safety is the highest priority in this study involving pregnant women and their infants. Ongoing eligibility will be assessed before each vaccination. The safety monitoring strategy includes active surveillance for immediate, solicited, and unsolicited adverse events through seven days after each investigational product administration, followed by passive surveillance thereafter. Pregnancy complications are monitored throughout the study period. Events meeting protocol-defined criteria are reported as adverse events of special interest (AESIs) or serious adverse events (SAEs), as appropriate. Infants undergo comprehensive clinical assessments at birth and at 6 weeks and 6 months of age. At the 6-month visit, neurodevelopment will be assessed using the Bayley Scales of Infant and Toddler Development™, Fourth Edition (Bayley™-4), to facilitate the early identification of developmental concerns and referral for further evaluation and care, where indicated. [33] Non-pregnant participants in the immunogenicity arm undergo urine pregnancy testing before each vaccination and receive counselling on pregnancy prevention.

Participants who withdraw from the study are not replaced. As part of participant safety, participants may have vaccination postponed or terminated in several scenarios. Vaccination will be postponed if a participant has received another vaccine such as tetanus toxoid (Td) within two weeks prior to the planned investigational product administration. Vaccination is also postponed if a participant has a fever (≥38.5°C axillary) or moderate or severe acute illness or infection on the day of investigational product administration. Participants may receive the study doses later upon resolution of these conditions. Vaccination will be discontinued if a participant experiences anaphylaxis or another severe reaction to the investigational product, or develops a clinically significant AE, SAE, or biological abnormality related to the previous investigational product administration that, in the investigator’s opinion, contraindicates further vaccination. Non-pregnant participants, or pregnant participants who have delivered, who have a positive urine pregnancy test after receiving at least one dose of Hecolin^®^ will discontinue further vaccination during the inadvertent pregnancy but will continue study follow-up, including safety assessments and HEV surveillance. Following delivery, participants who wish to complete the vaccination series will be offered the remaining Hecolin^®^ dose(s) through routine clinical practice. Participants who discontinue vaccination will be encouraged to continue all remaining study visits and assessments to ensure ongoing safety follow-up. Potential study-related risks also include blood collection, breast milk collection, and intramuscular injection. Blood collection may cause transient discomfort or vasovagal reactions, including fainting, which will be managed with appropriate supportive care by qualified medical personnel.

All SAEs will be recorded on the SAE reporting form and submitted to the sponsor within 24 hours of initial receipt of the information. The Principal Investigator will also report SAEs within seven days of initial receipt of the information to the Drug Regulatory Authority of Pakistan (DRAP), NBC and the site EC. All SAEs will be followed until resolution, stabilization, or until the investigator determines that the event has become chronic.

Study monitoring is conducted on behalf of the sponsor by the contract research organization (CRO), DRK Pharma Solutions, based in Pakistan. Planned blinded monitoring visits are performed throughout the study at the Aga Khan University Clinical Trial Unit, the four referral centres, and Kharadar Hospital. In addition, designated unblinded monitors conduct unblinded monitoring visits at the Clinical Trial Unit every four weeks to oversee study procedures requiring treatment allocation.

An internal Safety Monitoring Committee reviews blinded safety data for potential safety signals before Data and Safety Monitoring Board (DSMB) meetings and reports its findings to the Study Medical Monitor. The independent DSMB is composed of experts with relevant clinical, statistical, and ethical expertise who are independent of the sponsor and funder and have no competing interests related to the study. The DSMB periodically reviews accumulating safety data, including solicited and unsolicited adverse events, serious adverse events, adverse events of special interest, suspected unexpected serious adverse reactions, and adverse events leading to study withdrawal. Meetings include an open session, attended by the Study Medical Monitor and other designated study personnel, and a closed session attended only by DSMB members and unblinded statistician, during which unblinded safety data are reviewed. The DSMB also reviews study progress, participant recruitment, accrual and retention, and the overall benefit–risk profile of the trial, while considering relevant external scientific or therapeutic developments that may affect participant safety or trial conduct. Based on these reviews, the DSMB provides recommendations to the sponsor regarding continuation, modification, temporary suspension, or termination of the trial. The DSMB convenes at prespecified safety review milestones throughout the study, with additional ad hoc meetings convened if emerging safety concerns are identified.

To conceal allocation, central randomization is used with block sizes undisclosed to prevent identification of patterns. All study staff and participants are blinded to the arm assignment and only the study pharmacist and pre-defined staff (e.g., Independent statistician, unblinded clinical operations lead, randomization manager) are unblinded, are not involved in the safety assessment of participants, and are instructed not to comment on the experimental agent to blinded study staff. Any decision to unblind is made in concert between the Principal Investigator and the Medical Monitor. Emergency unblinding will only occur when knowledge of treatment allocation is essential for the participant’s clinical management, such as following a vaccine-related death or life-threatening SAE, when knowledge of treatment assignment may influence clinical decision-making or study management.

### Sample size

A total of 2,358 healthy women (2,208 pregnant women and 150 non-pregnant women) will be enrolled in the study. The 2,208 pregnant women will be stratified by gestational age stratified (1:1 between the second and third trimesters) and randomly assigned in a 1:1 ratio to receive either Hecolin^®^ (Arm 1) or placebo (Arm 2). Within each of Arms 1 and 2, 150 participants will be enrolled in the immunogenicity cohort. The 150 non-pregnant women (Arm 3) will receive Hecolin^®^ and serve as the non-pregnant comparator group for the immunogenicity analysis.

The sample size for evaluating and comparing the pregnancy-related safety events among recipients of two doses of Hecolin^®^ and placebo was calculated using a precision-based approach. The sample size calculation was based on the following assumptions:

- The anticipated event rate is 5% in both the Hecolin^®^ and placebo groups.
- The anticipated relative risk (RR; event rate in the Hecolin^®^ group divided by the event rate in the placebo group) is 1.0, indicating no difference between groups.
- The target precision for the estimated RR is a two-sided 95% confidence interval (CI) ranging from 0.67 to 1.50 (CI width = 0.83).
- The target precision for the event rate within each group is a two-sided 95% CI of 3.66% to 6.65% (CI width = 2.99%), assuming an observed event rate of 5%.
- An overall dropout rate of 20% is assumed.

The sample size was also intended to provide 90% power to detect at least one adverse event if the true event rate is 0.2% among pregnant women receiving Hecolin^®^. The overall study population of 2,358 participants (including pregnant and non-pregnant women and accounting for a 20% dropout rate) further increases the probability of observing infrequent adverse events occurring at a rate of approximately 0.2%.

Based on these assumptions, a sample size of 883 pregnant participants per group would provide the desired precision for both the estimated relative risk and the event rate. After allowing for a 20% dropout rate, a total of 1,104 pregnant participants per group (2,208 pregnant participants in total) will be enrolled.

For the primary immunogenicity outcomes, the sample size calculation was based on the following assumptions:

- The non-inferiority margin is 0.67-fold difference of geometric mean ratio (GMR) between the ratio of geometric mean concentration (GMC) in pregnant women receiving Hecolin^®^ and that in the non-pregnant women receiving Hecolin^®^.
- The coefficient of variation of GMC on the original scale is 1.5 (equivalently, the standard deviation of geometric mean titre (GMT) on the log 10 scale is 0.63) based on available data. [31]
- The true ratio of GMC on the original scale is 1.

Based on the above assumptions, 117 pregnant women and 117 non-pregnant women would provide 80% power at a one-sided 2.5% significance level to detect the non-inferiority of immunogenicity of Hecolin^®^ in pregnant women compared to non-pregnant women. After accounting for 20% dropout rate, the study requires 150 pregnant women and 150 non-pregnant women for primary immunogenicity endpoint analysis.

### Statistical analysis plan

Safety analyses will be performed for all participants who receive at least one dose of the investigational product (Safety Analysis Set). Safety endpoints include immediate adverse reactions (within 30 minutes), solicited AEs (within 7 days), unsolicited AEs (within 28 days after vaccination), pregnancy-related, neonatal and infant AESIs, and SAEs throughout the study period. Safety endpoints will be summarized descriptively, including the number and proportion of participants experiencing each endpoint together with corresponding two-sided Clopper–Pearson 95% confidence intervals (CIs). Solicited and unsolicited AEs will be summarized according to maximum severity and relationship to the investigational product. Adverse events will be coded using the latest version of the Medical Dictionary for Regulatory Activities (MedDRA) and summarized by system organ class and preferred term. Pregnancy-related AESIs, neonatal and infant AESIs will be summarized using the same approach, with between-group ratios and corresponding two-sided Score 95% CIs also presented.

Immunogenicity analyses will be performed using both the per-protocol set (PPS) and the full analysis set (FAS), with the PPS serving as the primary analysis population and the FAS used for sensitivity analyses. The FAS will include all participants who receive at least one dose of the investigational product and have at least one post-baseline immunogenicity assessment. The PPS will include all eligible randomized participants who receive planned vaccinations, have at least one immunogenicity assessment after respective vaccination, and have no major, important protocol deviations Immunogenicity analyses will be based on anti-HEV IgG geometric mean concentrations (GMCs), seroconversion rates (SCRs), geometric mean concentration ratios (GMRs), differences in SCRs, and their corresponding 95% confidence intervals (CIs) at each scheduled assessment time point. For the primary immunogenicity analysis, anti-HEV IgG GMCs measured 4 weeks after the second vaccination will be compared between pregnant Hecolin^®^ recipients (Arm 1) and non-pregnant Hecolin^®^ recipients (Arm 3). The GMR and its two-sided 95% CI will be calculated as the antilogarithm of the difference between the mean log-transformed antibody concentrations in Arms 1 and 3 using a general linear regression model, with adjustment for age and baseline imbalances, if necessary. Non-inferiority of immune responses in pregnant women will be concluded if the lower limit of the two-sided 95% CI for the GMR exceeds the prespecified non-inferiority margin of 0.67. The primary non-inferiority analysis will be conducted using a one-sided significance level of 2.5%. No adjustment for multiplicity will be applied to secondary analyses, and statistical significance for these analyses will be assessed using a two-sided significance level of 5%. Secondary and exploratory immunogenicity endpoints at other scheduled time points will be summarized descriptively.

Missing immunogenicity data will not be imputed. No interim analysis is planned.

#### Study data handling and protection

All study data will be entered into electronic case report forms within an electronic data capture system. Participant confidentiality will be protected through assignment of unique identifier codes that do not contain any personal identifiers (e.g., date of birth). Qualified study staff will enter data from source documents, which will be maintained in secure, access-restricted locations. Access to identifiable participant information will be restricted to authorized study personnel. Authorized representatives of the sponsor, contract research organization, ethics committees, and regulatory authorities may review identifiable source documents as required for study monitoring, auditing, or regulatory inspections. Automated range checks, edit checks, and predefined response options will be used to minimize data entry errors. Data queries will be generated regularly, and ongoing data cleaning will be performed throughout the study. No personal identifiers will be included in study reports or publications. Study records will be securely retained after study completion in accordance with institutional policies and applicable regulatory requirements.

#### Patient and public involvement

Prior to study initiation, the study design was shared with key national stakeholders in Pakistan, including members of the national obstetrics and gynecology society, the National Institute(s) of Health, and the Drug Regulatory Authority of Pakistan (DRAP). This engagement was undertaken to inform stakeholders of the planned study and to support the Investigational New Drug (IND) submission for its conduct in Karachi, Pakistan.

## Supporting information

Schedule of Procedures

## Data Availability

No datasets have been generated or analysed yet because this manuscript describes the protocol for an ongoing study.

## Ethics and dissemination

### Ethics approval

This study protocol has been reviewed and approved by the National Bioethics Committee (NBC) of Pakistan (Reference number: 4-87/NBC-910), the institutional Ethics Review Committee (ERC) of the Aga Khan University (Reference number: 8298), and the Institutional Review Board (IRB) of the International Vaccine Institute (IVI) (Reference number: 2022-007). The study is conducted in accordance with the principles of the Declaration of Helsinki and relevant regulations of the ICH Good Clinical Practice. All protocol amendments and SAEs are reported to and reviewed by DRAP and involved IRB and EC for approval and oversight of participant safety. The sponsor is the International Vaccine Institute. The study sponsor provides clinical trial insurance to cover the costs of medical care and compensation, where applicable, for study-related injuries. This trial is registered at the ClinicalTrials.gov (NCT05808166).

### Consent

Written informed consent is obtained from all participants. Participation in the trial is strictly voluntary, and participants may withdraw from the study at any time without affecting their access to routine medical care. Participants receive reimbursement for transportation costs and compensation for time spent attending study visits.

### Dissemination

Results will be published in peer-reviewed medical journals and presented at international immunization guideline committee meetings (e.g., Strategic Advisory Group of Experts on Immunization (SAGE)) and conferences. All publications (e.g., manuscripts, abstracts, oral/slide presentations and book chapters) from the study will be reviewed by each investigator prior to submission.

## Role of the sponsor, manufacturer and funders

The International Vaccine Institute (IVI) is the trial sponsor and designed the study, in collaboration with AKU investigators and external experts. IVI monitors trial conduct as part of oversight and will lead analysis and data dissemination. The funders (Gates Foundation, Coefficient Giving (formerly Open Philanthropy), Thrasher Research Fund) have no role in the design, conduct, or analysis of this study except that the Gates Foundation supported an external expert review of the protocol and Statistical Analysis Plan. Hecolin^®^ and the test kits for the immunogenicity assessments were donated by Innovax and Wantai, respectively. Innovax supported the preparation of the regulatory documents for the clinical trial application as a manufacturer.

## Author contribution

All authors contributed to protocol development and subsequent revisions. KRS, JL, LY, DRK, AR, and NT from the International Vaccine Institute, as the study sponsor, developed the initial study protocol. IN, AH, NA, SN, LS, and FJ reviewed the protocol and contributed to protocol revisions from the study site perspective. MB, KRT, and BLK contributed to the design and protocol development of the breast milk and infant serum analyses for the assessment of maternal antibody transfer. WC, XY, and RW reviewed the protocol and contributed to its development, particularly with respect to Hecolin^®^ and its relevant data from the manufacturer’s perspective. All authors critically reviewed the manuscript for important intellectual content, approved the final version for publication, and agreed to be accountable for all aspects of the work.

## Acknowledgement

This work was supported by grants from the Gates Foundation, Coefficient Giving, and the Thrasher Research Fund. We gratefully acknowledge their generous support, which made this study possible. The International Vaccine Institute gratefully acknowledges the partnership and financial contributions provided by the governments of the Republic of Korea, Sweden, Austria, Finland, India, Thailand, and the Philippines. We also sincerely thank the study teams for their dedication and contributions to the protocol development and updates, including Citra Vravita Khrisna, SengEn Her, Ju Yeon Park, Somyoung Cho, and Sujin Hwang (International Vaccine Institute, Seoul, Republic of Korea), as well as the study team members at Aga Khan University.

