## Supplementary material for "Safety and immunogenicity of recombinant hepatitis E vaccine in healthy pregnant women between 14 and 34 weeks of gestation and non-pregnant women of reproductive age: Protocol for a Phase II, randomized, observer-blinded, placebo-controlled trial": Schedule of Procedures

Table 1. Schedule of Study Procedures/Schedule of Events for pregnant and non-pregnant participants

| Visit Number | V1 | V2 | HV/ PC1[a] | V3 | HV/ PC2[a] | V4 | V5[f] | V6[b] | HV/ PC3[a] | V7 | V8 |
|---|---|---|---|---|---|---|---|---|---|---|---|
| Visit Day | -14 to 0 | 0 | 7 | 28 | 35 | 56 | Delivery | 168 | 175 | 196 | 336 |
| Visit Week (W) | -2 to 0 | 0 | 1 | 4 | 5 | 8 | - | 24 | 25 | 28 | 48 |
| Visit Month (M) | - | 0 | - | 1 | - | 2 | - | 6 | - | 7 | 12 |
| Visit Window | - | - | +3D | +7D | +3D | +7D | +7D[c] | +7D | +3D | +14D | ±7D |
| Approximate GA (W) | - | 14-34 | - | 18-38 | - | - | - | | | | |
| Screening | X | | | | | | | | | | |
| Informed Consent[d] | X | | | | | | | | | | |
| Eligibility Assessment | X | X | | X | | | | X | | | |
| Demographic Information | X | | | | | | | | | | |
| Height / Weight | X | | | | | | | X[f] | | | |
| Medical History | X | | | | | | | | | | |
| Prior/Concomitant Medication[e] | X | X | X | X | X | X | X | X | X | X | X |
| Obstetric History[f] | X | | | | | | | | | | |
| Prenatal Information[f] | X | X | | X | | X | | | | | |
| Delivery Information[f] | | | | | | | X | | | | |
| Pregnancy Confirmation and Dating (Ultrasound)[f] | X | | | | | | | | | | |
| Physical Examination[g] | X | X | | X | | X | X | X | | X | X |
| Vital Signs | X | X | | X | | X | X | X | | X | X |
| Fetal Heart Tone Monitoring[f] | X | X | | X | | X | | | | | |
| Urine Pregnancy Test[h] | X | X | | X | | | | X | | | |
| Pregnancy Prevention Counseling [h] | X | X | | X | | X | | X | | | |
| Enrollment and Randomization | | X | | | | | | | | | |
| Blood Sample: Immunogenicity[i] | | 8mL | | 8mL | | 8mL | 8mL | 8mL | | 8mL | 8mL |
| Blood Sample: Screening[j,k] | 15mL | | | | | | | | | | |
| Breast Milk Sample[i, l] | | | | | | | 10mL | | | 10mL | 10mL |
| IP Administration[m] | | X | | X | | | | X | | | |
| Adverse Events[n] | X | | | | | | | | | | |
| Immediate Adverse Events | | X | | X | | | | X | | | |
| Solicited Adverse Events[o] | | X | | X | | | | X | | | |
| Unsolicited Adverse Events[p] | | X | | | | | | X | | | |
| AESI[q] | | X | | | | | | | | | |
| SAE[r] | X | | | | | | | | | | |
| Pregnancy Reporting [h] | X | | | | | | | | | | |
| Diary Card (DC) Distribution | | DC1 | - | DC2 | - | DC3 | DC4 | DC5 | - | - | - |
| Diary Card (DC) Collection | | - | - | DC1 | - | DC2 | DC3 | DC4 | - | DC5 | - |
| Diary Card (DC) Review | | | X | X | X | X | X | X | X | X | |
| Study Completion Form | | | | | | | | | | | X |

a. HV/PC: Home visit or phone call. If a participant prefers, visits to site will also be allowed instead of HV/PC.

b. If the third dose (5 months interval between the second dose) comes before delivery, V6 will be adjusted to inject the IP within 14 days after delivery and the subsequent visit schedules, HV/PC3 (V6+1week), V7 (V6+4weeks) and V8 (V6+24weeks), will be changed accordingly with the indicated visit window maintained.

c. In case of home delivery or delivery at any other place than the study site, the procedures can be performed within 7 days window.
d. For pregnant participants, informed consent for both pregnant participant and their infant will be obtained.
e. After 28 days of each IP administration, concomitant medication history related to AESI and SAEs will only be recorded.
f. Will be performed in pregnant participants only.
g. Full physical examinations will be performed at screening and delivery (V5), and targeted physical examinations will be conducted during subsequent visits. For unscheduled visit, an attending physician will decide suitable physical examination method based on his/her judgement.
h. Will be performed in non-pregnant participants. For pregnant participants, if V6 is scheduled 6 weeks or later after delivery, a urine pregnancy test will be performed.
i. Samples for immunogenicity assessment will be taken from immunogenicity subsets (n=150) from each arm. Breast milk samples will be applicable for arms 1 and 2 only.
j. For those who have documented laboratory result at AKU lab from routine antenatal care within the last 14 days, overlapping laboratory assessment will not be repeated during screening visit. If missed or abnormal results are detected, the applicable lab can be repeated as a part of screening assessment. For ABO/Rh, if the test was performed at AKU lab within the current pregnancy for the pregnant participants, the test does not need to be repeated during screening visit.
k. Laboratory assessment includes complete blood count (CBC) with differential, chemistry (Sodium, Potassium, Chloride, Calcium, Phosphate, ALT/AST, ALP, total bilirubin, GGT, glucose, BUN/Creatinine), Serologic tests (HIV antibody test, anti-HCV antibody, HBsAg for all participants/Syphilis test for pregnant participant only), ABO/Rh for pregnant participant only, Urine analysis with microscopic examination (protein, glucose, RBC), Urine pregnancy test for non-pregnant participants only.
l. Breast milk sample will be collected at delivery, 6 weeks and 6 months after delivery from immunogenicity subset in arms 1 and 2 only.
m. Participants receiving placebo will be provided with the 3 doses of Hecolin® or other locally available adult vaccines after the completion of infant visit to ensure equal benefit as the test vaccine group.
n. Adverse events will be collected from initiation of screening to before vaccination.
o. Solicited adverse events will be collected for 7 days following the IP administration. The same duration will be applicable for solicited adverse events related to unscheduled visit.
p. Unsolicited adverse events will be collected for 28 days following the IP administration. The same duration will be applicable for unsolicited adverse events related to unscheduled visit.
q. AESI: Adverse event of special interest. Pregnancy-related AESI will be collected during pregnancy in pregnant participants only. The same duration for the event collection/reporting will be applicable for AESI related to unscheduled visit.
r. SAE: Serious adverse event. SAEs will be collected throughout the study period.

- As per the study objectives, the intervals and windows of the visits will be counted based on each dose vaccination.

### Table 2. Study Procedures/Schedule of Events for infant participants

| Visit Number | | V1 | V2 | V3 |
|---|---|---|---|---|
| **Visit Weeks** | | **Delivery [a]** | **6 weeks** | **24 weeks** |
| **Visit Window** | | **-** | **± 14D** | **± 14D** |
| **Infant Enrollment ID Assignment[b]** | | X | | |
| **Document APGAR Score, Gestational Age at Birth[c]** | | X | | |
| **Gross Anomaly Check** | | X | | |
| **Demographic Information** | | X | | |
| **Weight / Length / Head Circumference** | | X | X | X |
| **Medical History[d]** | | X | | |
| **Concomitant Medication[e]** | | X | X | X |
| **Physical Examination[f]** | | X | X | X |
| **Vital Signs** | | X | X | X |
| **Developmental Assessment[g]** | | | | X[k] |
| **Blood Sample for Immunogenicity[h]** | **Cord Blood[i]** | 10 ml | | |
| | **Venous Blood** | | 3ml | 3ml |
| **Safety Follow Up[j]** | | X | | |
| **Study Completion Form** | | | | X |

a. In case of home delivery, procedures can be conducted within 7 days window.
b. The participant card will be given to the parent/guardian of the infant participant at the delivery visit.
c. APGAR score measured at 1, 5 and 10 minutes will be collected.
d. Any medical history during fetal period (e.g. anomaly detected by ultrasonography, fetal treatment conducted).
e. Concomitant medication related to AE/AESI/SAE will be collected.

f. Full physical examinations will be performed at all scheduled visits. For unscheduled visit, an attending physician will decide suitable physical examination method based on his/her judgment.
g. Bayley Scales of Infant and Toddler Development$^{TM}$, fourth edition (Bayley$^{TM}$-4) will be used for developmental assessment. (5)
h. Samples for immunogenicity assessment will be taken from infants born from pregnant immunogenicity subsets (n=150) of arm 1 and 2.
i. An infant blood draw should be preferentially obtained within 24 hours of birth if cord blood is not collected at delivery but is permissive up to 7 days post-delivery.
j. Any AEs reported in infants will be categorized as unsolicited adverse events. AESI and SAE will be collected throughout the study period.
k. In case an infant is delivered preterm;
- All infant participants do not belong to immunogenicity subsets, will undergo V2 (6 weeks after delivery) based on chronological age, and V3 (6 months after delivery) based on adjusted age.

- Infants belonging to immunogenicity subsets, will undergo V2 (6 weeks after delivery) based on chronological age and V3 (6 months after delivery), also based on chronological age. If the gap between chronological and adjusted ages exceeds the window period of 14 days, an additional visit (V3') should be scheduled at an adjusted age of 6 months. Should this additional visit (V3') be necessary, developmental assessment and study completion form will be postponed from V3 (6 months by chronological age) to V3' (6 months by adjusted age). At the additional visit (V3') scheduled at 6 months by adjusted age, all study procedures for V3 will be conducted, excluding blood sampling.

Figure 1. Visit Schedules

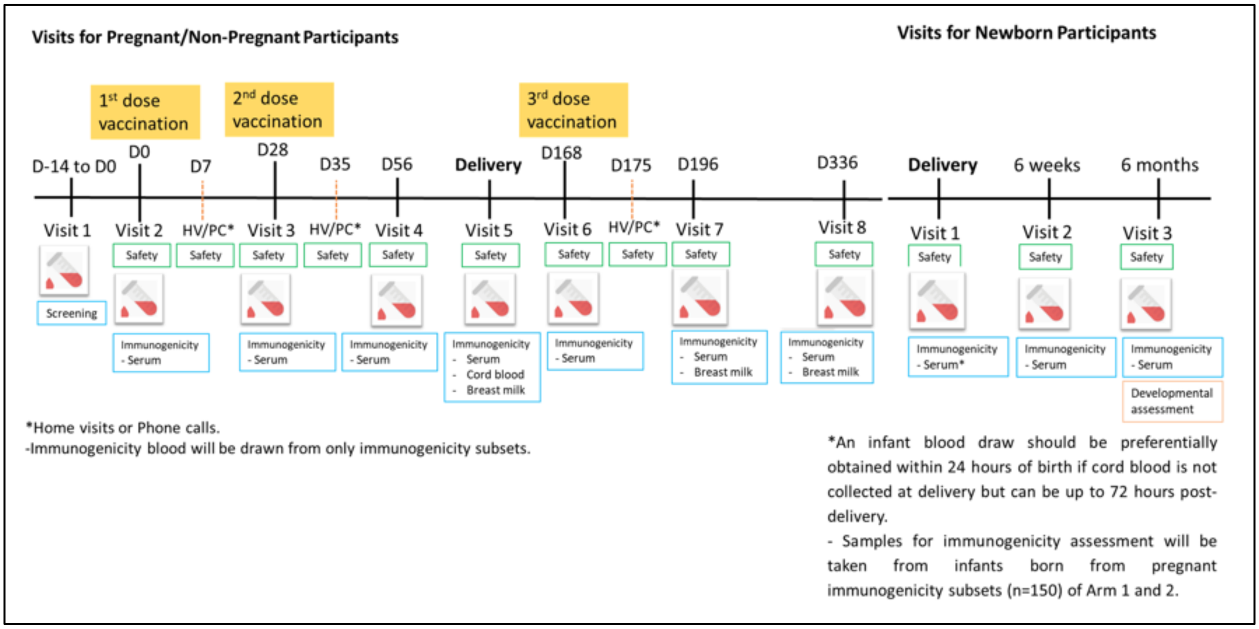
